# The Challenge of Using Epidemiological Case Count Data: The Example of Confirmed COVID-19 Cases and the Weather

**DOI:** 10.1101/2020.05.21.20108803

**Authors:** Francois Cohen, Moritz Schwarz, Sihan Li, Yangsiyu Lu, Anant Jani

## Abstract

The publicly available data on COVID-19 cases provides an opportunity to better understand this new disease. However, strong attention needs to be paid to the limitations of the data to avoid making inaccurate conclusions. This article, which focuses on the relationship between the weather and COVID-19, raises the concern that the same factors influencing the spread of the disease might also affect the number of tests performed and who gets tested. For example, weather conditions impact the prevalence of respiratory diseases with symptoms similar to COVID-19, and this will likely influence the number of tests performed. This general limitation could severely undermine any similar analysis using existing COVID-19 data or similar epidemiological data, which could, therefore, mislead decision-makers on questions of great policy relevance.

**One Sentence Summary:** Measurement issues in the currently available data on confirmed COVID-19 cases undermine the analyses of the drivers of the spread of the disease.

## Main text

The publicly available datasets on confirmed COVID-19^v^ cases and deaths provide a key opportunity to better understand the drivers of the pandemic. Research using these datasets has been growing at a very fast pace (see an indicative list of references in supplementary material 1). However, little attention has been paid to the reliability of this type of epidemiological data to make statistical inferences.

Our initial aim was to produce a detailed statistical analysis of the relationship between weather conditions and the spread of COVID-19. This question has attracted significant attention from the media (e.g. 1, 2) and the research community (e.g. 3, 4; see a wider list in supplementary material 1) due to the possibility that summer weather might slow the spread of the virus. After going through all the steps of such an analysis, we reached the unexpected conclusion that the limitations of the available COVID-19 data are so severe that we would not be able to make any reliable statistical inference. This applies, for example, to the data provided by the John Hopkins University (5) and the data collated by Xu et al. (2020) (6).

This is a concerning yet very important finding considering that such data is being widely used to make crucial policy decisions on a wide range of topics. Since invalid causal inferences could be made with the publicly available COVID-19 data, and then enter policy-making discourse, there is an urgent need to raise awareness among the scientific community and decision-makers regarding the limitations of the information at their disposal. The elements discussed in this paper are also likely to be applicable to other epidemiological datasets obtained with insufficient testing and monitoring, either during exceptional epidemics or seasonal outbreaks.

Several challenges could undermine any causal statistical analysis of the influence of a potential determinant, such as the weather, on the spread of COVID-19. To start, confounding variables are likely to pose a significant problem: many factors (e.g. changes in policy or social interactions) are simultaneously influencing how the disease spreads.

In addition, significant challenges come from the limitations of the COVID-19 case count data itself. Firstly, testing capacity has been a major issue in most countries. Before March 1^st^, 2020, very few countries had sufficient testing capacity. By April 30^th^, 2020, high-income countries had significantly increased their testing capacity, but testing remained critically infrequent in most low-and middle-income countries.^iv^ **Figure 1, panel a** illustrates the effect that insufficient testing capacity has on the number of confirmed cases. It distinguishes between three phases of limited (I), intermediate (II) and widespread (III) testing. In Phases I and II, there is a risk that the number of confirmed cases depends more on the number of tests available than on the actual number of people who have COVID-19, questioning the validity of any analysis relying too heavily on this data.

**Figure 1:**
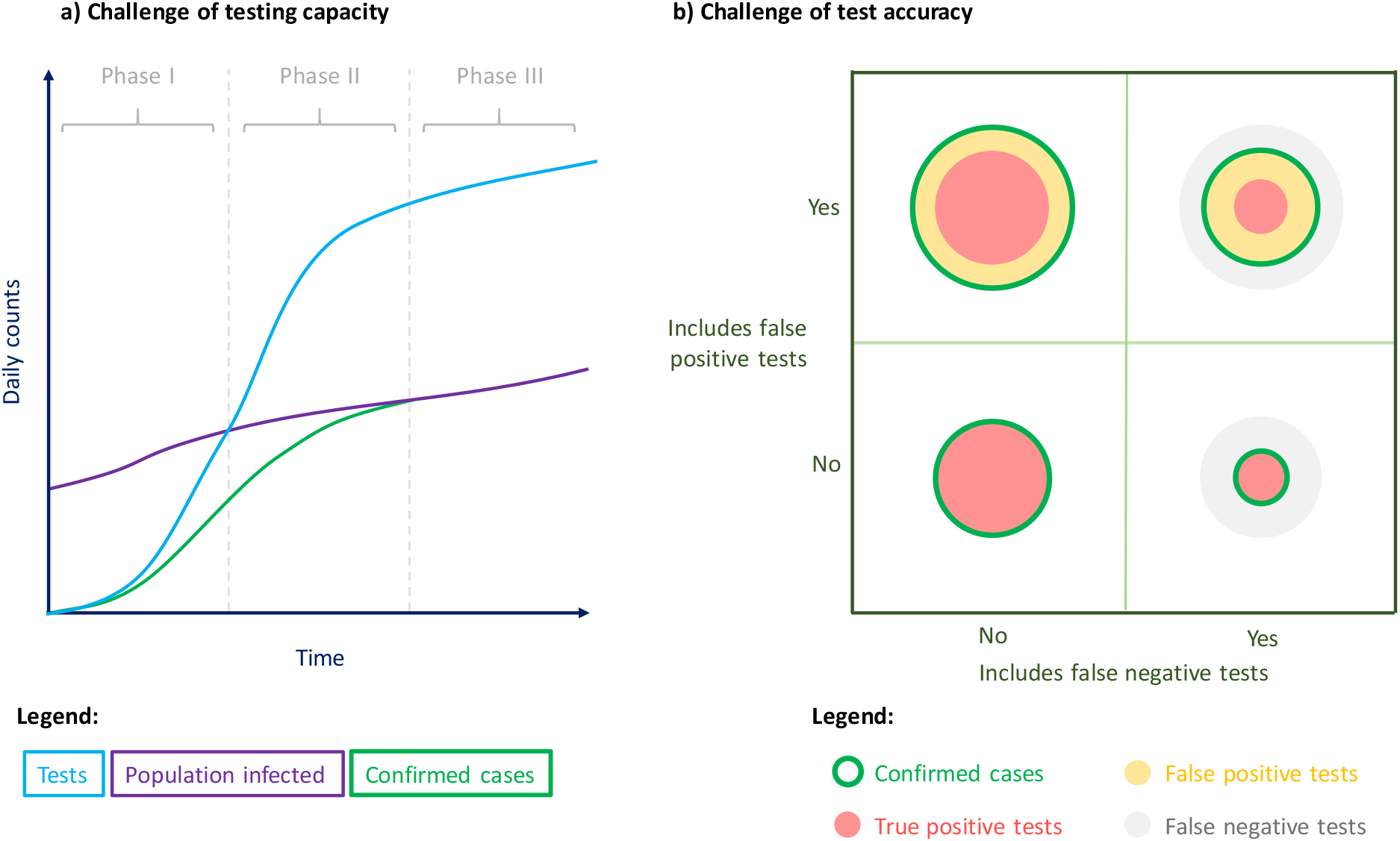
Difference between actual COVID-19 cases in population and reported confirmed COVID-19 cases. Confirmed COVID-19 cases (green) represent the number of people tested with a positive test result. They include false positive and exclude false negative tests. The circles of panel b represent the size of the populations with true positive, false negative or false positive tests. Quantities in the y-axis of panel a, as well as the size of the circles in panel b, do not represent any true value or proportion.

Moreover, there have been numerous concerns regarding the accuracy of the COVID-19 tests performed so far (7, 8, 9, 10). **Figure 1, panel b** illustrates the effects of both false-negative and false-positive test results on the number of confirmed cases. False-negative results would imply that the number of confirmed COVID-19 cases is underestimated. False-positive results would imply that people who do not have COVID-19 are included in the number of confirmed COVID-19 cases. Concerns regarding test accuracy create an additional problem of measurement that might affect statistical analyses.

The two above-mentioned challenges are inherent to all current datasets of COVID-19 confirmed case count and mortality. In addition, specific datasets may have imperfect geographical or time coverage.

To look at the impact of the weather on the spread of COVID-19, we initially used a well-established approach, similar to the ones used previously to look at the impact of the weather on other diseases (e.g. 11, 12) (see details in supplementary material 2). However, the fundamental measurement issues associated with the COVID-19 case count data cannot be corrected by statistical techniques, as we outline below.

The main problem is that the weather could be influencing the number of tests carried out and the segment of the population tested. For example, other respiratory diseases are often similar to COVID-19 in their symptoms (e.g. 13) and are more common during cold weather (e.g. 11, 12), which could influence the number of tests performed on people displaying symptoms of respiratory infection. Therefore, even if the model correctly identified the impact of the weather on COVID-19 case counts, it could not distinguish between the impact of the weather on the spread of the disease and its impact on testing. **Table 1** provides a non-exhaustive list of elements that could undermine any analysis of the impact of the weather on the spread of COVID-19 using data on confirmed cases. The evidence suggests that the weather may correlate with the number of tests conducted and who gets tested. We have not been able to find any specific COVID-19 related evidence that the weather would correlate with test accuracy (e.g. the weather affecting the nasopharyngeal or oropharyngeal swabs used in the PCR analysis), even though this could be possible.

**Table 1:**
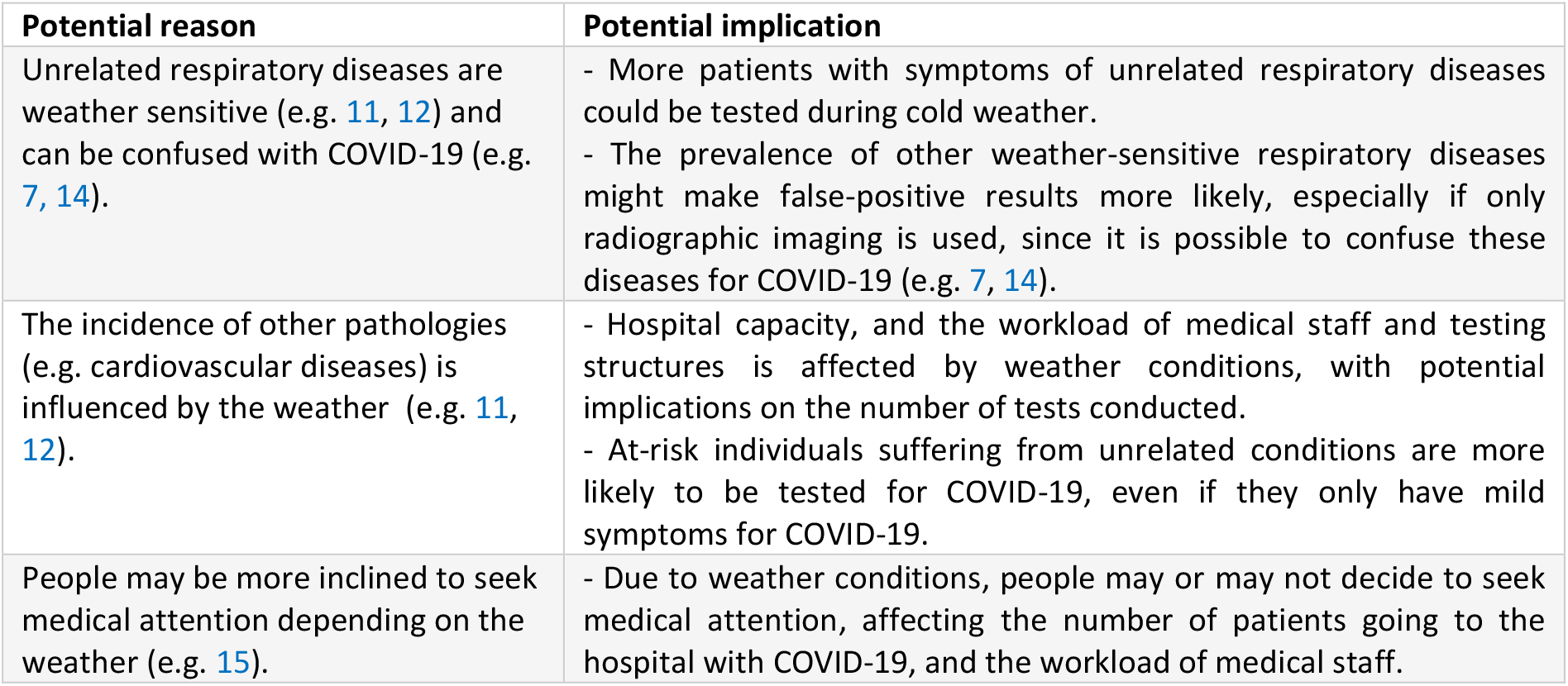
Non-exhaustive list of reasons why weather conditions could affect the number of COVID-19 tests carried out and who gets tested.

Other points of concern include: the fact that there may be indirect effects of weather conditions on other factors that could have an impact on the spread of COVID-19 (such as social interactions or air pollution); the heterogeneity of impacts across populations and subgroups within a population; or the fact that some people may have travelled and therefore been infected in a different place from where the cases are reported.

We ran our model (as detailed in the supplementary material 2) and provide results and robustness checks in supplementary material 3. The model would technically suggest a negative correlation (e.g. colder days would be associated with more confirmed COVID-19 cases, and hotter days with fewer cases). Yet, these results could be highly misleading since these estimates are likely to be substantially biased because of the aforementioned reasons.

**Figure 2, panel a**, provides an illustration of how we could have obtained a negative correlation even if temperature had no impact or a positive impact on the spread of COVID-19 in our sample. The total number of estimated cases is given by the size of the circles as a function of temperature (x-axis). The circles in green correspond to the effects we are interested in – those that explain the influence of temperature on the spread of COVID-19. If temperature has no effect on the spread of COVID-19, then the green circles should be the same size at low and high temperatures. The pink circles represent the possible effect of temperature on testing (as reported in **Table 1**) under the illustrative assumption that high temperatures reduce testing frequency. In this case, the overall result is a negative correlation between temperature and confirmed COVID-19 cases, even if temperature has no effect on the spread of the disease. In practice, we naturally do not know the direction of the bias caused by the effect of temperature on testing when using standard statistical methods. There is also no way for us to evaluate the contribution of each of these effects (green or pink) in our estimate. We arrive at the final size of the circles and cannot be sure if the association that we are interested in is either negative, null or positive.

**Figure 2:**
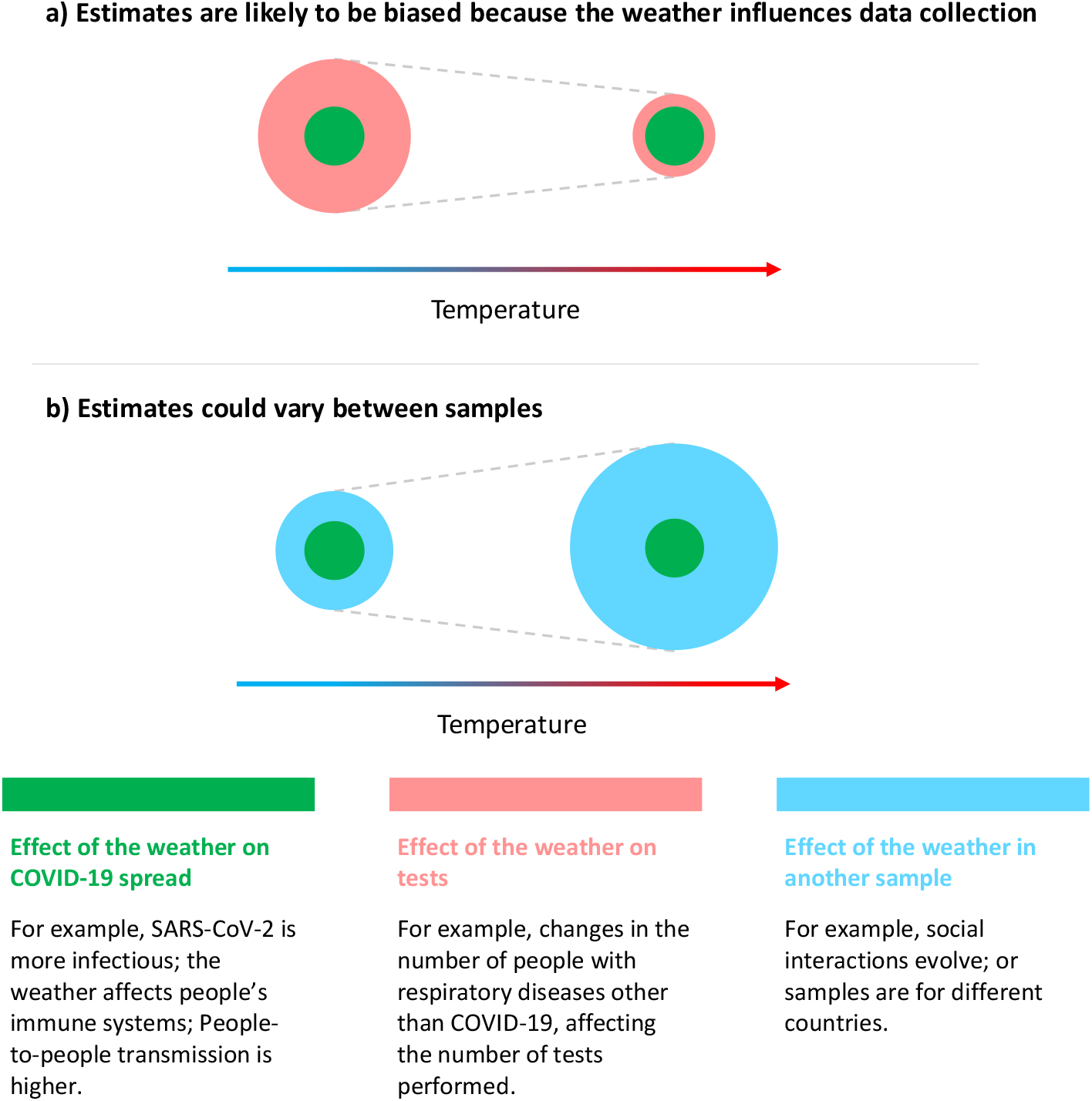
Effects potentially captured by our estimate. The size of the circles represents the estimated number of cases at different temperatures. These are examples that do not correspond to actual data. In these examples, we assume no correlation between temperature and the effects in green (see legend below), a negative correlation with the effects in pink (example 1) and a positive correlation with those in blue.

**Figure 2, panel b**, focuses on the risk that effects could be different across different samples. The circles in blue capture other underlying factors that are influenced by temperature (such as acclimatisation or the level of social interactions in the population), as well as other socioeconomic factors (such as the demographic characteristics of a population). These factors could be radically different in different regions but may also evolve over time (e.g. between winter and summer seasons).

There are strong reasons to be concerned with the scenario illustrated in **Figure 2, panel b**. In our sample, for example, we only have data from the start of the pandemic until end of April 2020; some countries (e.g. China) may be over-represented in the dataset; and the average daily temperature is relatively low at 10.5°C. Furthermore, many countries have implemented a stringent containment policy during the period covered by the sample. Containment policies may have heightened (or lowered) the sensitivity of the spread of the disease to the weather because social interactions are limited. We are not able to observe how the impact of the weather on COVID-19 might change at different gradients of social interaction. Finally, our estimate is based on small, observed changes in temperatures, and not on radical increases or reductions in temperatures. The spread of COVID-19 may respond differently to large variations in temperature, e.g. by 5°C or 10°C across seasons, making seasonal predictions even more unreliable.

Strong precautions need to be taken before using COVID-19 case count datasets for inference. The results of our model using existing COVID-19 data would seemingly imply a negative association between temperature and confirmed COVID-19 cases. Any projection of COVID-19 cases with such estimates could conclude that, during the upcoming months of June to September 2020, Southern Hemisphere countries would be exposed to higher risks of COVID-19 spread, and Northern Hemisphere countries to lower risks.^vii^ These types of unsubstantiated results could be used as a misinformed justification for an early relaxation of effective social distancing measures in the Northern Hemisphere.

These findings have equally strong implications for statistical analyses focusing on other questions that rely on COVID-19 confirmed case count and/or mortality count data. Even though the exact nature of the effects may change, such studies are also at risk of capturing the effect that their parameters of interest have on tests and test results. For example, studies interested in the effect of containment policies may have to consider that these policies substantially affect testing because they change the awareness of the disease in the population, political demands for more testing or the risk of contracting other respiratory diseases. Other studies may also produce estimates that are very specific to the current circumstances in the development of the pandemic and are, therefore, not suitable to use for forecasts of what could happen in the coming months.

In the medium term, more reliable data needs to be gathered, for example through experimental studies that randomly test a sample of the population for COVID-19. In the short term, we are in a situation of fundamental uncertainty about how different factors affect or are affected by the widespread societal changes we see with the COVID-19 pandemic. Therefore, scientists, policymakers, journalists and the general public need to be very cautious when discussing how the spread of COVID-19 correlates with the weather or any other factor.

In the long term, this paper suggests that more attention should be given to how epidemiological data is recorded and used during exceptional epidemics and seasonal outbreaks, since insufficient testing and monitoring can undermine essential statistical analyses. This article calls for the complementary use of different methods for data collection, such as random testing in samples of the population.

## Data Availability

All data and software are publicly available at https://github.com/moritzpschwarz/COVID-19-weather-Oxford.

## Acknowledgements.

For useful comments, we thank, among others, Eric Beinhocker, Cristina Castro, Antoine Dechezlepretre, Doyne Farmer, Matthieu Glachant, Jeongmin Thomas Han, David Hendry, Cameron Hepburn, José García Quevedo, Daniel Montolio, James Parkhurst, Rafael Perera, Felix Pretis, Andra Tartakowsky, Elisabet Viladecans, Judit Vall Castelo and the team of the Future of Cooling programme at the Oxford Martin School. For technical IT assistance, we thank David Ford. For copyediting, we thank Jack Smith.

## Authors contributions

Cohen is the first author. He had the original idea, wrote most of the paper and the code to produce the econometric analysis. He also coordinated the team. Jani ensured the material was consistent with the epidemiological evidence. Li produced the required climate data for the statistical analysis. Lu helped on literature review, on coding the econometric analysis and on producing the tables. Schwarz helped on data coding and matching and created the projections. All authors contributed to the text.

## Data and materials availability

all data and software are publicly available at https://github.com/moritzpschwarz/COVID-19-weather-Oxford.

## Supplementary Materials

1. References from the emerging literature on the spread of COVID-19
2. Data and methods to correlate weather conditions to confirmed COVID-19 cases
3. Main results and robustness checks
4. Information on projections

Table A1 – A6

Fig A1 – A2

## Footnotes

v In this article we follow Xu et al. (2020) who define COVID-19 cases as individuals for whom SARS-CoV-2 has been detected using rt-PCR.

vi Figures on testing are available in Our World In Data (accessed on May 1st, 2020): https://ourworldindata.org/coronavirus

vii We performed such a projection to confirm this point (see **supplementary material 4**).

